# Influencing factors of Anti-SARS-CoV-2-Spike IgG antibody titres in healthcare workers – A cross-section study

**DOI:** 10.1101/2022.05.10.22274912

**Authors:** Julia Reusch, Isabell Wagenhäuser, Alexander Gabel, Annika Eggestein, Anna Höhn, Thiên-Trí Lâm, Anna Frey, Alexandra Schubert-Unkmeir, Lars Dölken, Stefan Frantz, Oliver Kurzai, Ulrich Vogel, Manuel Krone, Nils Petri

## Abstract

**Background:** Against the background of the current COVID-19 infection dynamics with the rapid spread of SARS-CoV-2 variants of concern (VOC), above all the Omicron VOC, the immunity of healthcare workers (HCWs) against SARS-CoV-2 continues to be of high importance. Vaccination plays a central role in reducing the severity and potentially the spread of the disease. In healthcare, this is important to prevent disease-related staff shortages. However, there is a lack of data on factors influencing the humoral immune response.

**Aim:** The aim of our study was to determine factors influencing the level of Anti-SARS-CoV-2-Spike IgG after SARS-CoV-2 infection or vaccination in healthcare workers.

**Methods:** 1,750 study participants were recruited who met the following inclusion criteria: age ≥ 18 years, PCR-confirmed SARS-CoV-2 infection and/or at least one dose of COVID-19 vaccination, working in health care. Anti-SARS-CoV-2-Spike IgG titres were determined by SERION ELISA *agile* SARS-CoV-2 IgG.

**Results:** Mean Anti-SARS-CoV-2-Spike IgG levels increased significantly with the number of COVID-19 vaccinations (92.2 BAU/ml for single dose, 140.9 BAU/ml for two doses and 1,144.3 BAU/ml after threefold vaccination). Hybrid COVID-19 immunized respondents (after infection and vaccination) had significantly higher antibody titres compared with participants after infection only (525.4 BAU/ml vs. 105.7 BAU/ml). Anti-SARS-CoV-2-Spike IgG titres declined significantly with time after administration of the second vaccine dose. Smoking and high age were associated with lower titres.

**Conclusion:** Both recovered and vaccinated HCWs presented a predominantly good humoral immune response with decreasing antibody levels over the temporal course. Smoking and higher age limited the humoral SARS-CoV-2 immunity. This reduced immune response is an important aspect as people with these risk factors are recognized as people with an increased risk for a severe course of disease.

## 1. Introduction

Against the background of the ongoing COVID-19 pandemic [1] and the current infection dynamics with the rapid spread of the Omicron SARS-CoV-2 variant of concern (VOC) as well as high incidence levels [2, 3], the immunity of healthcare workers (HCWs) against SARS-CoV-2 continues to play a critical role in preventing disease-related staff shortages and keeping up public health care capacities [4-6]. COVID-19 vaccines have evolved as a key prevention strategy to reduce the severity of disease and combat the global spread of SARS-CoV-2 [7].

The humoral immune response against SARS-CoV-2 is investigated in order to provide forecasts regarding immunity and protection against severe courses of disease [8]. The low number of studies published to date show a significant correlation between neutralizing antibody titres and preservation from symptomatic SARS-CoV-2 infections [9]. The available data is still insufficient to make any concrete statements on the influencing factors of antibody titres considering the large number of possible combinations of COVID-19 vaccinations and/or SARS-CoV-2 infection.

Previously published studies on humoral Anti-SARS-CoV-2-Spike antibodies have been conducted in small cohorts or for only short observation periods without consideration of demographic factors, quality of life as well as ability to work. There is a lack of comparable real-life large-scale seroprevalence data, particularly in HCWs [10-13].

This study examines the seroprevalence of Anti-SARS-CoV-2-Spike IgG following SARS-CoV-2 infection and/or COVID-19 vaccination in HCWs and determines factors influencing antibody titres as a cross-section study.

## 2. Methods

### 2.1 Study setting

The data presented is part of the prospective CoVacSer cohort study, which examines SARS-CoV-2 immunity derived from serial blood samples as well as survey-based quality of life and ability to work in HCWs after COVID-19 vaccination and/or SARS-CoV-2 infection.

The CoVacSer study participants had to meet the following inclusion criteria: (i) age ≥ 18 years, (ii) signed consent form, (iii) 14 days minimal interval after first polymerase chain reaction (PCR) derived confirmation of SARS-CoV-2 infection and/or at least one dose of COVID-19 vaccination independent of the vaccination concept, and (iv) employment in the healthcare sector.

Serum blood samples for Anti-SARS-CoV-2-Spike IgG determination were collected combined with pseudonymised CoVacSer study surveys including demographic data, physical condition and personal risk factors in addition to World Health Organization Quality of Life (WHOQOL-BREF) [14, 15] and Work Ability Index (WAI) questionnaire [16].

Only serum blood samples that were accompanied by a signed consent form as well as a fully completed digital questionnaire have been taken into account for the data analysis. Following pseudonymisation, the matching of blood sample and survey was mediated based on date of birth and dates of SARS-CoV-2 infection or COVID-19 vaccination.

The arriving date of the serum was documented and equated with the specimen collection date. Blood samples with a gap of less than 14 days to the last SARS-CoV-2 infection or dose of COVID-19 vaccination were not regarded for evaluation in accordance with the study’s inclusion criteria.

Participants with vaccines that were not authorized by the European Medicines Agency (EMA) were excluded from this study. The following vaccines have been included due to EMA authorization throughout the data collection period: (i) BNT162b2mRNA (Comirnaty, BioNTech/Pfizer, Mainz/Germany, New York/USA), (ii) mRNA-1273 (Spikevax, Moderna, Cambridge/USA), (iii) ChAdOx1-S (VaxZevria, AstraZeneca, Cambridge/GB), (iv) Ad26.COV2-S (COVID-19 vaccine Janssen, New Brunswick/USA) [17].

The data presented in this study describes the cross-sectional seroprevalence of Anti-SARS-CoV-2-Spike IgG titres among HCWs after COVID-19 vaccination and/or SARS-CoV-2 infection at the time of study inclusion.

### 2.2 Data collection

The data collection period ranged from the 29^th^ of September 2021 to the 12^th^ of November 2021 during the fourth wave of the COVID-19 pandemic in Germany [3]. The federal vaccination campaign in Germany started on 27^th^ December 2020 with the consequent expansion of vaccination capacities [18]. Due to the initial vaccine shortage, vaccination was carried out according to a tiered plan, with HCWs assigned to the highest priority level [19].

Mainly HCWs from a single tertiary care hospital participated in the study, but HCWs from surrounding hospitals and private practice were also included in this study.

The questionnaire survey including WHOQOL-BREF [14, 15] and WAI [16] was performed using REDCap (Research Electronic Data Capture, projectredcap.org).

### 2.3 SARS-CoV-2 IgG ELISA

Anti-SARS-CoV-2-Spike IgG titres were determined by SERION ELISA *agile* SARS-CoV-2 IgG (SERION Diagnostics, Wuerzburg, Germany), technically carried out as an enzyme linked immunoassay (ELISA). This assay was selected as it has proven to be superior to comparable SARS-CoV-2 IgG ELISA [20] methods, having a high correlation with the neutralization titres [21-23].

The extinction values were photometrically measured operating with the Dynex Opsys MR™ Microplate Reader and Relevation Quick Link (Dynex technologies, Chantilly VA, USA) at 405 nm wavelength. The extinction was transferred to the manufacturer specific Serion IgG units per ml (U/ml) using the software easyANALYSE (SERION Diagnostics). These units were converted into the internationally established unit Binding Antibody Units per ml (BAU/ml) using the factor 2.1 according to the manufacturer’s instructions [24].

The threshold IgG values in the selected SERION assay were defined as < 10.0 U/ml (21.0 BAU/ml) for negative, ≥ 10.0 U/ml (21.0 BAU/ml) to < 15.0 U/ml (31.5 BAU/ml) for results at the borderline and ≥ 15.0 U/ml (31.5 BAU/ml) as positive. These values were chosen according to manufacturer’s instruction and IgG values above the threshold of 31.5 BAU/ml indicate at least a moderate neutralization capacity [21]. For detecting Anti-SARS-CoV-2-Spike IgG antibody levels beyond the maximum limit of 250 U/ml (525.0 BAU/ml), serum blood samples were diluted based on a dilution series with dilution factors both 10 and 100. Consequently, the measurement range of SERION ELISA *agile* SARS-CoV-2 IgG could be expanded.

### 2.4 Ethical approval

The study protocol was approved by the Ethics committee of the University of Wuerzburg in accordance with the Declaration of Helsinki (file no. 79/21).

### 2.5 Statistics

The statistical analyses were performed with the statistical programming language R (version 3.1.2) [25].

Statistical differences between the age distributions of male and female HCWs were separately calculated with the Kolmogorov Smirnov test against the corresponding age distribution within the German population in 2019 [26].

The analysis of Anti-SARS-CoV-2-Spike IgG titres was performed on logarithmized IgG titres (*Supplementary Figure 2*).

To analyse the effect of physical conditions and personal risk factors on logarithmized Anti-SARS-CoV-2-Spike IgG titres, a lasso regression was performed to identify factors that are associated with IgG (*Supplementary Figure* 4). This regression model included the factors age, gender, BMI, smoking, immune deficiency, drug intake, time between serum sampling and last SARS-CoV-2 immunizing event (infection or vaccination), vaccination concept and other lifestyle parameters obtained from the study questionnaire. Thereby, the factor of age was binarized due to the categories 18 to 40 years old (18 -40) and older than 40 years (41+). Using a tenfold cross-validation procedure, the model parameters of the lasso regression model were estimated and the model having the lowest mean squared error (MSE) of ∼0.62 was chosen (*Supplementary Figure 5*). An advantage of the lasso regression is the shrinkage of factors down to zero if they do not show any relevant association to the dependent variable. Thus, the factors age, gender, BMI, profession, immune deficiency, smoking, satisfaction with the own health status, medical treatment needs, having a meaningful life, concept of vaccination, and time until serum sampling showed associations to the corresponding Anti-SARS-CoV-2-Spike IgG titres.

Modelling the differences of IgG titres with respect to the applied vaccination concept (*Figure 3*), age, gender, BMI, smoking, immune deficiency, time between serum sampling and last SARS-CoV-2 immunizing event and further associated factors (*Supplementary Figure 7*) defined by the lasso regression, a multiple linear regression model was applied to the data. Estimating the coefficients of this model based on a data set with unequal sample sizes, a generalised least square fit was performed with the R package *nlme* [27, 28].

Based on the estimated coefficients from the regression model, statistical differences between subgroups of the categorical variables (*Figure 3, Figures 4 A -D, Supplementary Figure 4*) were calculated using the marginal estimated means. The statistically significance of the pairwise differences were calculated with the Tukey statistics. The post hoc pairwise comparisons were performed by using the *emmeans* package [29]. To correct against multiple testing, the resulting p values were adjusted using the Benjamini-Yekutielie procedure [30]. Adjusted p values below a significance level of 0.05 were considered statistically significant.

The reproducible script of all statistical analyses can be accessed at https://github.com/AlexGa/Influencing-factors-of-Anti-SARS-CoV-2-Spike-IgG-antibody-titres-in-healthcare-workers.

## 3. Results

### 3.1. Specimen collection and participant recruitment

From the 29^th^ of September 2021 to the 12^th^ of November 2021, 1,782 study participants were recruited, who submitted a serum blood sample and completed the CoVacSer study survey. Out of 1,782 persons, 1,750 (98.2%) participants were finally included. 32 persons did not meet the inclusion criteria: 10 neither had a PCR confirmed SARS-CoV-2 infection nor received at least one dose of a COVID-19 vaccine. 12 submitted a blood sample before the defined minimal interval of 14 days to the PCR confirmation of the latest SARS-CoV-2 infection and/or the recent administration of a COVID-19 vaccination dose. 10 persons provided insufficient information on type or pattern of COVID-19 vaccination in the CoVacSer study survey (*Figure 1*).

**Figure 1:**
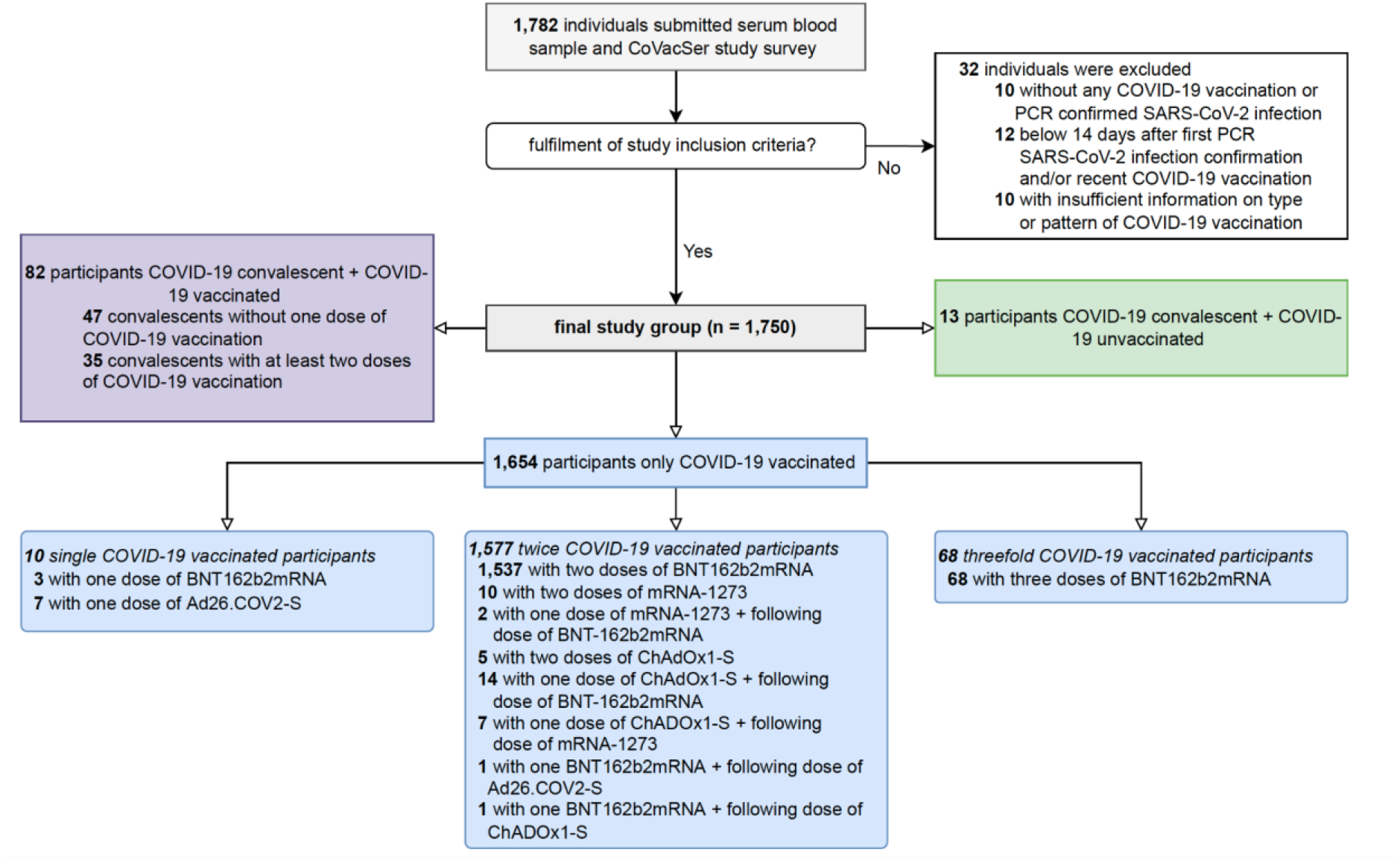
Enrolment of study participants and subjects’ characteristics concerning possible combinations of SARS-CoV-2 immunization. PCR: Polymerase Chain Reaction

Out of the final study group, 13 (0.7%) participants were SARS-CoV-2 infection convalescent but not vaccinated against COVID-19, 82 (4.7%) had a hybrid SARS-CoV-2 immunity (SARS-CoV-2 infection convalescents with at least one COVID-19 vaccination). 1,577 (90.1%) were twice COVID-19 vaccinated, 68 (3.9%) threefold vaccinated. Among the twice COVID-19 vaccinated participants, two consequent doses of BNT162b2mRNA were administered in 1,537 cases (87.8%), 10 (0.6%) respondents were vaccinated twice with mRNA-1273 and 5 (0.3%) have received two doses of ChAdOx1-S. 14 (0.8%) participants were vaccinated with a first dose of ChAdOx1-S followed by a second dose of BNT162b2mRNA, 7 (0.4%) followed by a second dose of mRNA-1273. 2 (0.1%) respondents were COVID-19 vaccinated with one dose of mRNA-1273 followed by one dose of BNT162b2mRNA, 1 (0.1%) respondent each received a dose of BNT162b2mRNA followed by a dose of ChAdOx1-S and Ad26.COV2-S, respectively. 7 (0.4%) study participants received a single dose of Ad26.COV2-S.

The median interval between the first and the second dose of BNT162b2mRNA was 21 days (IQR: 21-21), in case of double mRNA-1273 administration 38 days (IQR: 28-42) and in case of twice ChAdOx1-S 77 days (IQR: 66-91). For heterologous vaccination schedules with one dose of ChAdOx1-S followed by one dose of BNT162b2mRNA, the median vaccination interval was 77 days (IQR: 70-82), in case of ChAdOx1-S followed by mRNA-1273 84 days (IQR: 80-84).

### 3.2 Study population

1,410 out of the 1,750 participants assigned themselves to the female gender (80.6%), 340 to the male gender (19.4%). The age of enrolled study participants ranged from 18 to 75 years (median: 39, IQR 30-52), median age of the female participants was 40 years (IQR: 29-53) and 38 years in male participants (IQR: 31-49, *Figure 2*). In total, 626 (35.8%) of the participants worked as nurses, of which 541 (30.9% of all) were female and 85 (4.9%) were male. 322 (18.4%) were physicians, of which 190 (10.9%) were female and 132 (7.5%) were male. Further professional groups involved other HCWs with contact to patients (22.2% in total) and HCWs without patient contact (23.7% in total).

**Figure 2:**
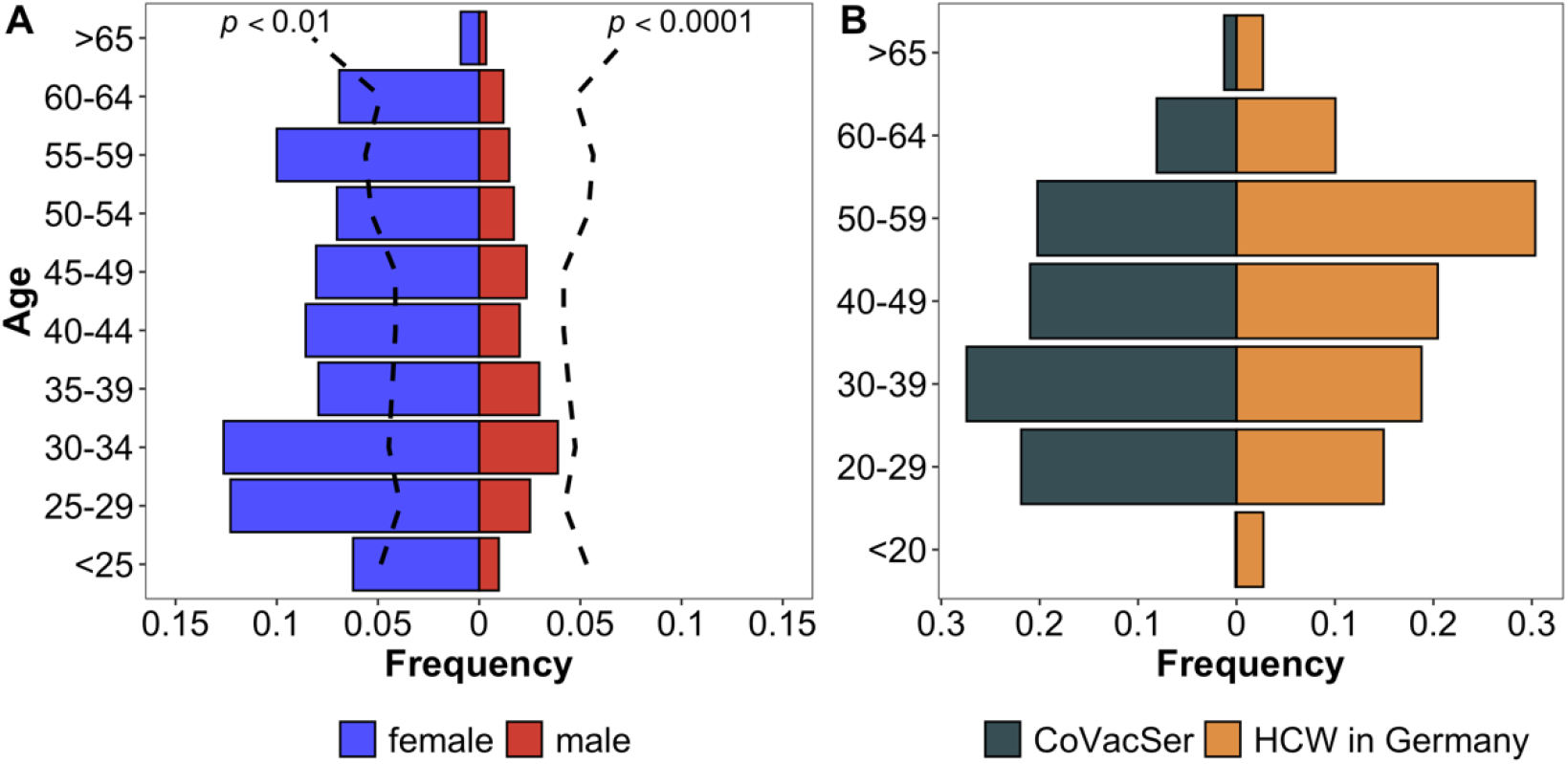
Characterization of study population compared to the German general public and HCWs in Germany. Comparison of CoVacSer study population to reference populations. **Fig. 2A** portrays the enrolled HCW study population (portrayed in gender separated blue and red bars, n = 1,750) in comparison to the demographic composition of the German general public considering gender and age (black broken line, Kolmogorov-Smirnov-Test) as of December 31, 2020 [26]. **Fig. 2B** compares the age structure in 10-year categories as a percentage of respondents included in the study (blue bars) with the total number of HCWs in Germany (red bars) [31]. Data Source: German federal office for statistics, German federal health reporting [26, 31, 32].

### 3.3 Anti-SARS-CoV-2-Spike IgG titres depending on vaccination and infection

The collected serum specimens from the enrolled HCWs contained an Anti-SARS-CoV-2-Spike IgG range from 6.3 to 6,517.8 BAU/ml (geometric mean: 161.4 BAU/ml, IQR: 201.6 BAU/ml). Among participants administered with one dose of the COVID-19 vaccines, Anti-SARS-CoV-2-Spike IgG titres ranged from 15.2 to 402.1 BAU/ml (geometric mean: 92.2 BAU/ml, IQR: 69.8 BAU/ml), with 80.0% (8/10) reaching the positive IgG threshold of 31.5 BAU/ml. In case of double COVID-19 vaccination, Anti-SARS-CoV-2-Spike IgG levels between < 6.3 to 4,794.7 BAU/ml (geometric mean: 140.9 BAU/ml, IQR: 173.5 BAU/ml) were obtained, with 95.2% (1,502/1,573) exceeding 31.5 BAU/ml. All study participants that received three or more doses of COVID-19 vaccination had titres beyond the threshold (68/68, range 79.5 to 6,227.8 BAU/ml, geometric mean: 1,114.4 BAU/ml, IQR: 1,312.2 BAU/ml).

In SARS-CoV-2 recovered and COVID-19 unvaccinated participants, the obtained Anti-SARS-CoV-2-Spike IgG levels ranged from 22.8 to 299.8 BAU/ml (geometric mean: 105.8 BAU/ml, IQR: 127.7 BAU/ml), 92.3% (12/13) showed IgG levels above 31.5 BAU/ml. Among the hybrid COVID-19 immunized respondents, the IgG levels between 15.5 to 6,517.8 BAU/ml (geometric mean: 525.4 BAU/ml, IQR: 656.9 BAU/ml) were detected, with 98.8% (81/82) above 31.5 BAU/ml (*Figure 3, Supplementary Table 1*).

**Figure 3:**
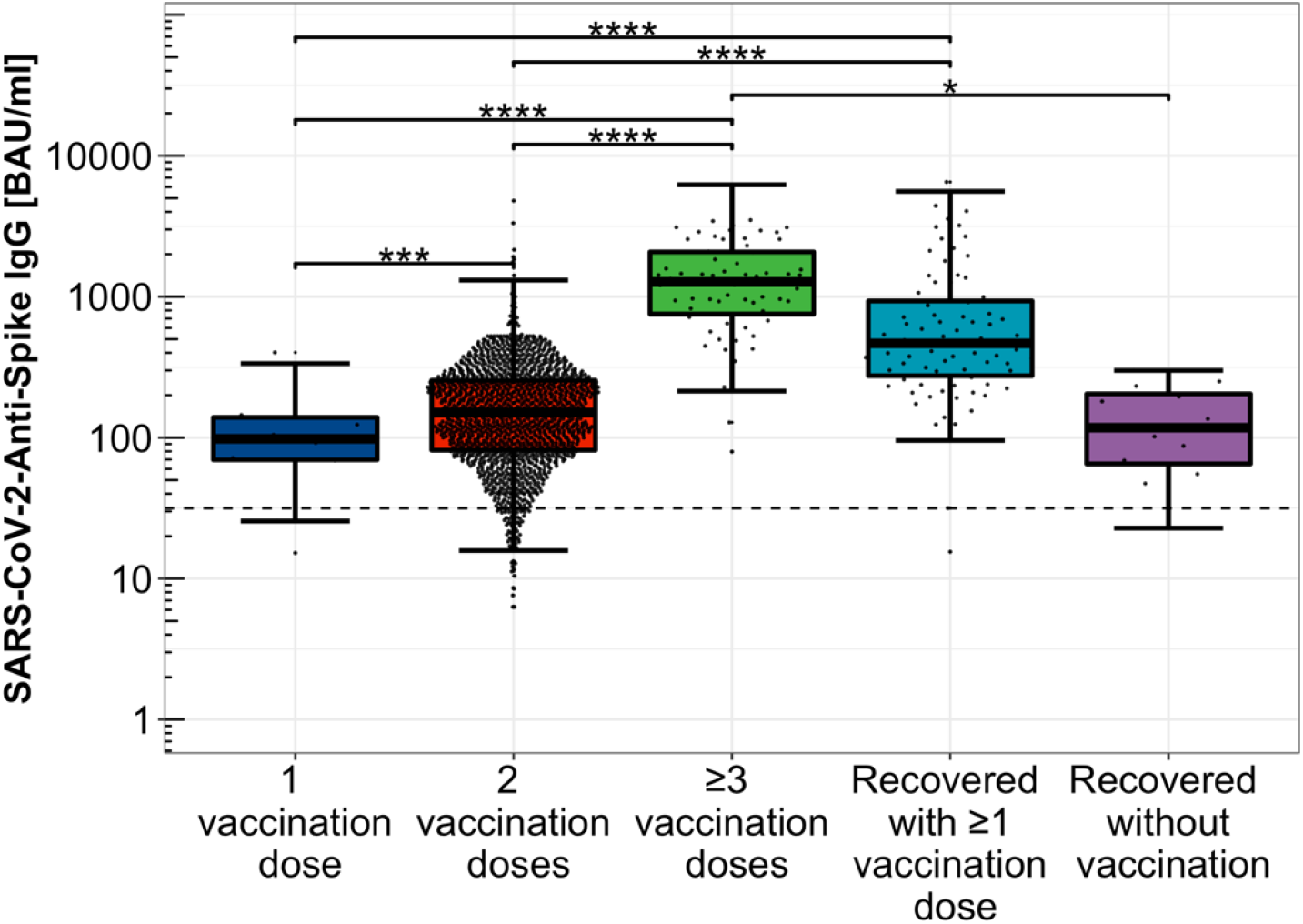
Distribution of Anti-SARS-CoV-2-Spike IgG levels depending on immunization scheme. Distribution of Anti-SARS-CoV-2-Spike IgG titres among single, double, and threefold COVID-19 vaccinated participants, only COVID-19 convalescent study participants as well as hybrid immunized participants including SARS-CoV-2 infection convalescence and COVID-19 vaccination, logarithmically scaled. ****: *p* < 0.0001 ***: *p* < 0.001 **: *p* < 0.01 *: *p* < 0.05 *BAU/ml: Binding Antibody Units per millilitre*

The pairwise differences in SARS-CoV-2-Anti-Spike IgG titres with respect to different vaccination concepts were statistically significant when comparing one with two administered doses of COVID-19 vaccination (p < 0.001), dual versus at least threefold vaccination (p < 0.0001), convalescents without any COVID-19 vaccination compared to threefold or more vaccinated participants (p < 0.05), and comparing the hybrid immunized with both single and double COVID-19 vaccinated participants (p < 0.0001).

### 3.4 Course of time of SARS-CoV-2 Anti-Spike IgG levels after last immunizing event

The time interval from the last SARS-CoV-2 immunizing event to the moment of study participation ranged from 14 to 569 days (geometric mean: 197.6 days, IQR: 77 days). Among the convalescent study participants, 168 to 569 days (geometric mean: 267 days, IQR: 96 days) passed since the first PCR confirmation of the latest SARS-CoV-2 infection. With respect to statistical outliers, one HCW with a time interval of 569 days between SARS-CoV-2 infection and study participation and without additional immunization was removed from further statistical analysis (*Supplementary Figure 3*). Within the hybrid COVID-19 immunized subgroup, the time since last event ranged from 14 to 292 days (geometric mean: 115 days, IQR: 114 days). In case of one administered dose of COVID-19 vaccination, 14 to 211 days had passed (geometric mean: 59 days, IQR: 73 days), among the double COVID-19 vaccinated participants 21 to 304 days (geometric mean: 221 days, IQR: 66 days), and 14 to 127 days (geometric mean: 33 days, IQR: 30 days) in case of a threefold COVID-19 vaccination.

A strong association between time since last immunizing event and SARS-CoV-2-Spike IgG titres could be detected (*Figure 4E*). The IgG titres are negatively correlated (c = -0.45) and show a significant decrease over time (p < 0.0001).

**Figure 4:**
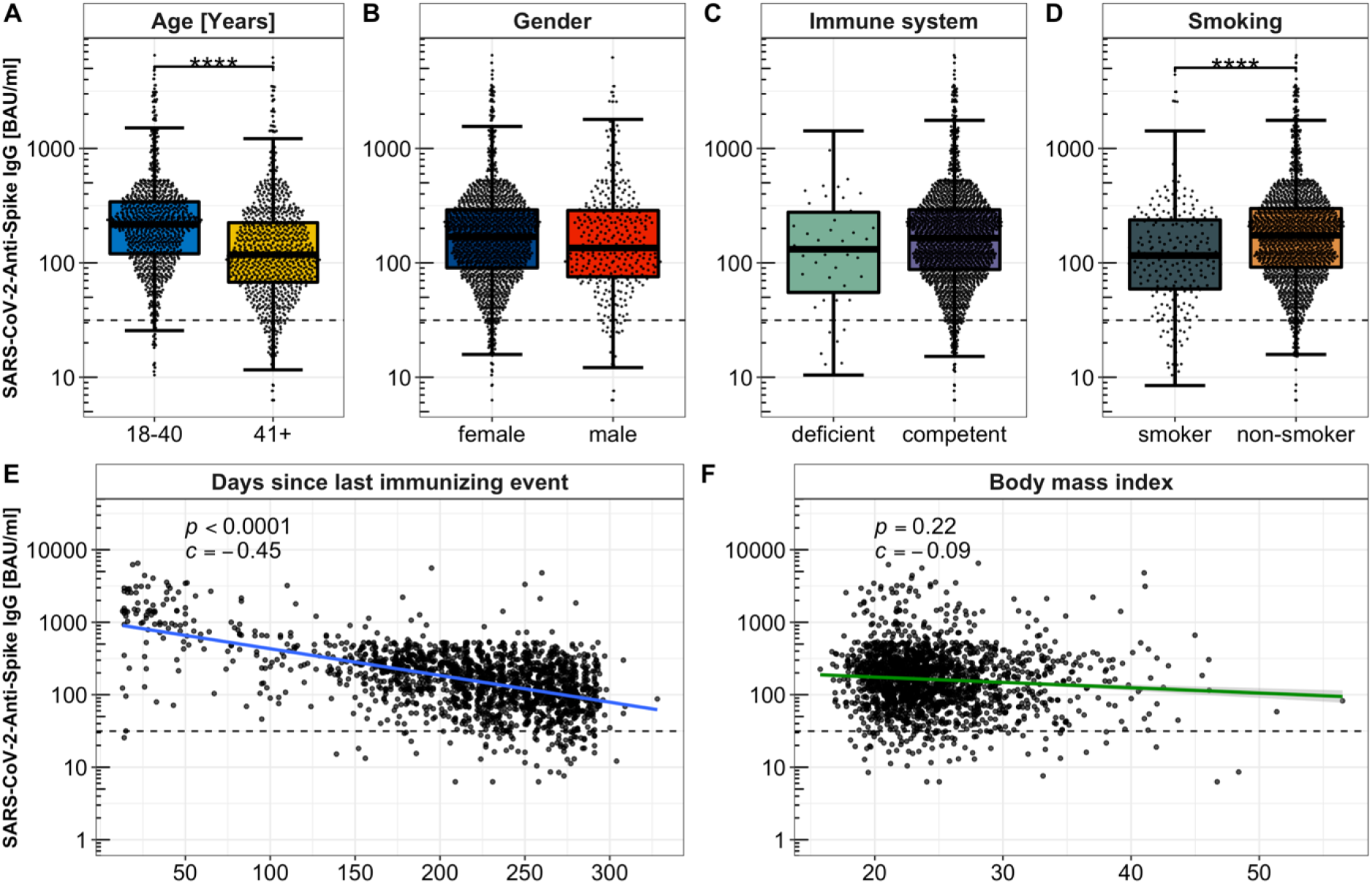
Anti-SARS-CoV-2-Spike IgG levels depending on individual physical properties. Fig. **4A**: Comparison of Anti-SARS-CoV-2-Spike IgG levels depending on age in the categories 18 -40 years versus older than 40 years. Fig. **4B**: Anti-SARS-CoV-2-Spike IgG titres among smokers versus non-smokers. Fig. **4C**: Anti-SARS-CoV-2-Spike IgG levels of immune deficient compared to immune competent respondents. Fig. **4D**: Anti-SARS-CoV-2-Spike IgG titres depending on sex. Fig. **4E:** Chronological decline of Anti-SARS-CoV-2-Spike IgG levels among double vaccinated respondents, logarithmically scaled, each dot represents a study participant. Fig. **4F**: Anti-SARS-CoV-2-Spike IgG titres depending on BMI logarithmically scaled. BAU/ml: Binding Antibody Units per millilitre *\*\*\*\*: p <* 0.0001

### 3.5 Identification of Anti-SARS-CoV-2-Spike IgG titres influencing factors

Based on the lasso regression model, the following variables were selected to be associated with Anti-SARS-CoV-2-Spike IgG levels: age group, gender, BMI, field of employment, immune deficiency, smoking, contentedness with personal health, personal dependency of medical treatment, subjective usefulness of life, contact to SARS-CoV-2 infected patients, vaccination concept as well as passed time after the last SARS-CoV-2 immunizing event.

The following variables were not associated to the Anti-SARS-CoV-2-Spike IgG titres: general contact to patients, respondent subjective evaluation as COVID-19 risk patient, life quality, long-term medication, subjective feeling of safety as well as subjective enjoyment of life (*Supplementary Figure 4 and 5*).

### 3.6 Influence of individual factors on Anti-SARS-CoV-2-Spike IgG titres

Anti-SARS-CoV-2-Spike IgG levels were significantly lower among study participants aged 40 years or older (939/1,750, 53.7%, mean: 119.8 BAU/ml, IQR: 148.5 BAU/ml), compared to the younger participants (811/1,750, 46.3%, mean: 201.2 BAU/ml, IQR: 214.5 BAU/ml, (p < 0.0001, *Figure 4A*).

In the male group, the mean Anti-SARS-CoV-2-Spike IgG titre was 146.1 BAU/ml (IQR: 206.4 BAU/ml), among female participants 161.1 BAU/ml (IQR: 194.1 BAU/ml) implementing slightly higher antibody levels in the female subgroup (p > 0.05, *Figure 4B*).

2.6% (45/1,750) reported an illness accompanied by an immune deficiency with an obtained mean Anti-SARS-CoV-2-Spike IgG titre of 120.0 BAU/ml (IQR: 225.5 BAU/ml) in this subgroup. Comparison of immune deficient to immune competent respondents (mean Anti-SARS-CoV-2-Spike IgG level: 159.2 BAU/ml, IQR: 195.6 BAU/ml) regarding antibody levels did not show statistically significant differences. (p > 0.05, *Figure 4C*).

222 (12.7%) of the enrolled study participants were smokers with an average level of 11.4 pack-years (py), which is defined as the number of cigarette packages smoked per day multiplied by years of smoking per person. The geometric mean of Anti-SARS-CoV-2-Spike IgG levels in case of smoking was 112.8 BAU/ml (IQR: 179.3 BAU/ml), among the non-smokers as 166.1 BAU/ml (IQR: 203.2 BAU/ml), portraying smoking as a statistically significant restriction of Anti-SARS-CoV-2-Spike IgG levels (p < 0.0001, *Figure 4D*).

The median body weight was 68 kg (IQR: 60 to 80 kg), with median gender-specific values of 65 kg (IQR: 58 to 75 kg) in the female and 80 kg (IQR: 73 to 90 kg) in the male subgroup. Combined with the reported body size, with a median of 169 centimetres (IQR: 164 to 175 cm, median of 168 cm (IQR: 163 to 172 cm) in the female and 180 cm (IQR: 176 to 185 cm) in the male subpopulation), an average body-mass index of 24 kg/m^2^ was calculated (median of 23 kg/m^2^ among the female, 25 kg/m^2^ among the male study participants. Anti-SARS-CoV-2-Spike IgG levels show very weak associations to BMI, spearman correlation of -0.09 (*Figure 4F*).

## 4. Discussion

Overall, SARS-CoV-2 convalescent as well as COVID-19 vaccinated and hybrid immunized HCWs presented Anti-SARS-CoV-2-Spike levels indicating at least a moderate neutralizing capacity [21]. Mean Anti-SARS-CoV-2-Spike IgG titres increased with the number of administered doses of COVID-19 vaccines (92.2 BAU/ml for single, 140.9 BAU/ml for double and up to 1,144.4 BAU/ml after threefold vaccination). Study participants after SARS-CoV-2 infection and without a COVID-19 vaccination had a mean Anti-SARS-CoV-2-Spike antibody titre of 105.8 BAU/ml. In comparison, SARS-CoV-2 convalescents with at least a single vaccination (hybrid immunized) had a mean titre of 525.4 BAU/ml, which confirms the importance of the COVID-19 vaccination as an additional contribution to proper humoral protection against SARS-CoV-2 after infection [33-35]. The antibody levels in case of a completed basic immunization, comprising two doses of COVID-19 vaccination, were significantly lower than in the hybrid immunized group. This cross-section study highlights the relevance of the COVID-19 vaccination as a prevention measure, especially in the critical cohort of HCWs that are highly exposed to SARS-CoV-2.

Anti-SARS-CoV-2-Spike IgG levels were each statistically significantly lower in smokers and older participants than in the respective comparison groups. Further investigations are needed to assess the impact of nicotine consumption in more detail, such as the influence of the number of pack-years on Anti-SARS-CoV-2-Spike IgG levels. COVID-19 vaccine dose adjustment could provide better humoral protection for smokers who are already recognized as people with an increased risk for a severe course of disease [36]. Further, a trend towards less, but not statistically significant impairment of the humoral immune response to COVID-19 vaccination and/or SARS-CoV-2 infection was observed among participants suffering from immune deficiency. More precise information regarding the extent and type of the immune suppression and its influence on the humoral immunity is necessary to be able to offer specific and individualized COVID-19 vaccination recommendations for the future. A restricted humoral SARS-CoV-2 immune response of the older participants is a further risk factor for a SARS-CoV-2 infection and severe course of disease. This underlines the importance of a COVID-19 vaccination as a prevention strategy, especially in older HCWs.

However, the significance of the factors that have a negative impact on Anti-SARS-CoV-2-Spike IgG levels is still unclear, as threshold values for IgG titres that protect against infection or a severe course are still lacking. Comparability with international research is restricted due to data being presented that is not converted to the global standardized Anti-SARS-CoV-2-Spike IgG of BAU/ml following the WHO recommendations in a proportion of other studies [24].

The obtained Anti-SARS-CoV-2-Spike IgG titres showed a significant decrease over time after the last SARS-CoV-2 immunizing event indicating a declining humoral immune response after the baseline immunization with COVID-19 vaccines. This examination represents a cross-section of data at the beginning of a prospective surveillance study of HCWs that analyzes the humoral immune response as well as quality of life and ability to work.

The data presented should be interpreted considering the possible influence of the following limitations. Firstly, the study population consists of 80.5% female HCWs and thereby represents the typical female focused gender composition in the public healthcare sector in Germany with a female share of 75.5% among HCWs in 2019 [32]. Nevertheless, due to the large study population and the gender differentiated data analysis, this only slightly limits the transferability of the described findings to both HCWs and the public. Secondly, the vaccines administered are heterogeneously distributed in our cohort, with BNT162b2mRNA double administration accounting for the largest proportion by far. The share of Anti-SARS-CoV-2-Spike IgG titres after vaccination with other COVID-19 vaccines than BNT162b2mRNA is consequently limited and needs to be investigated in further studies. Similarly, the intervals between the individual COVID-19 vaccine administrations vary as recommended vaccination intervals are not adhered to in all cases. This also limits the generalisability but represents a real-life scenario. Another limitation resides within unknown, not PCR confirmed SARS-CoV-2 infections, which might lead to higher Anti-SARS-CoV-2-Spike IgG levels after vaccination. Only Anti-SARS-CoV-2-Spike IgG were serologically obtained, consequently the differentiation of antibody levels being solely a result of COVID-19 vaccination or consequences of an unknown SARS-CoV-2 infection is not possible. However, unknown SARS-CoV-2 infections among HCWs might be seen as less frequent in comparison to the general public due to a strict set of measures to reduce and prevent the spread of SARS-CoV-2 in healthcare institutions including regularly implemented employee testing strategies [5]. All subject-related data, except for the objective serological Anti-SARS-CoV-2-Spike IgG measurement, were collected by the means of an electronic questionnaire. Consequently, the obtained data is influenced by the subjectivity of the study participants. We tried to counteract this by using the standardized questionnaires WHOQOL-BREF [14, 15] and WAI [16]. Further confounding aspects regarding the Anti-SARS-CoV-2-Spike IgG seroprevalence, which are not queried in the survey, cannot be denied. Because of pseudonymisation and data privacy of the study participants, query of the exact medication including agent and dose of immune suppression was relinquished. This limits the interpretation of the Anti-SARS-CoV-2-Spike IgG levels of HCWs with immune deficiency and its consequences on the immune response after COVID-19 vaccination.

Based on the presented findings, further examinations regarding the temporal course of Anti-SARS-CoV-2-Spike IgG levels and the influence of further COVID-19 vaccine administrations or SARS-CoV-2 infections are urgently needed. In addition, research on correlation of titre values and protection against infection and/or a severe course of disease should be intensified.

## 5. Conclusion

The humoral immunity against SARS-CoV-2 within the examined cohort of HCWs presents as predominantly good among both convalescent and COVID-19 vaccinated participants. The significantly higher Anti-SARS-CoV-2-Spike IgG levels in the hybrid immunized subgroup compared with convalescent-only participants highlight the importance of an additional vaccination after convalescence. Further, significantly higher titres of Anti-SARS-CoV-2-Spike IgG were observed after the third vaccination compared with only twice vaccinated participants. The temporal course of the titre levels will be monitored in the ongoing CoVacSer study.

The reduced humoral immune response in smokers and older health care workers adds to the already recognised increased risk for a severe course of disease in these groups of individuals. This is especially important among the highly exposed cohort of HCWs.

The correlation of breakthrough infections to Anti-SARS-CoV-2-Spike IgG levels needs to be further investigated, especially in the context of the Omicron VOC and possible future SARS-CoV-2 VOCs.

## Data Availability

Additional data that underlie the results reported in this article, after de-identification (text, tables, figures, and appendices) as well as the study protocol, statistical analysis plan, and analytic code is made available to researchers who provide a methodologically sound proposal to achieve aims in the approved proposal on request to the corresponding author.

https://github.com/AlexGa/Influencing-factors-of-Anti-SARS-CoV-2-Spike-IgG-antibody-titres-in-healthcare-workers

## 6. Author contributions

All authors had unlimited access to all data. Julia Reusch, Isabell Wagenhäuser, Alexander Gabel, Manuel Krone, and Nils Petri take responsibility for the integrity of the data and the accuracy of the data analysis.

Conception and design: Thiên-Trí Lâm, Anna Frey, Alexandra Schubert-Unkmeir, Lars Dölken, Stefan Frantz, Oliver Kurzai, Ulrich Vogel, Manuel Krone, Nils Petri.

Anti-SARS-CoV-2-Spike IgG titre determination: Julia Reusch, Isabell Wagenhäuser.

Trial management: Julia Reusch, Isabell Wagenhäuser, Annika Eggestein, Anna Höhn, Manuel Krone, Nils Petri.

Statistical analysis: Julia Reusch, Isabell Wagenhäuser, Alexander Gabel, Manuel Krone, Nils Petri.

Obtained funding: Oliver Kurzai, Ulrich Vogel.

First draft of the manuscript: Julia Reusch, Isabell Wagenhäuser, Alexander Gabel, Manuel Krone, Nils Petri.

Reviewing and modifying the manuscript and approving its final version: Annika Eggestein, Anna Höhn, Thiên-Trí Lâm, Anna Frey, Alexandra Schubert-Unkmeir, Lars Dölken, Stefan Frantz, Oliver Kurzai, Ulrich Vogel.

## 10. Acknowledgements

We explicitly thank the serological diagnostic laboratory staff for sharing their laboratory, and for all their help and advice.

## Supplementary material

**Supplementary Fig. 1:**
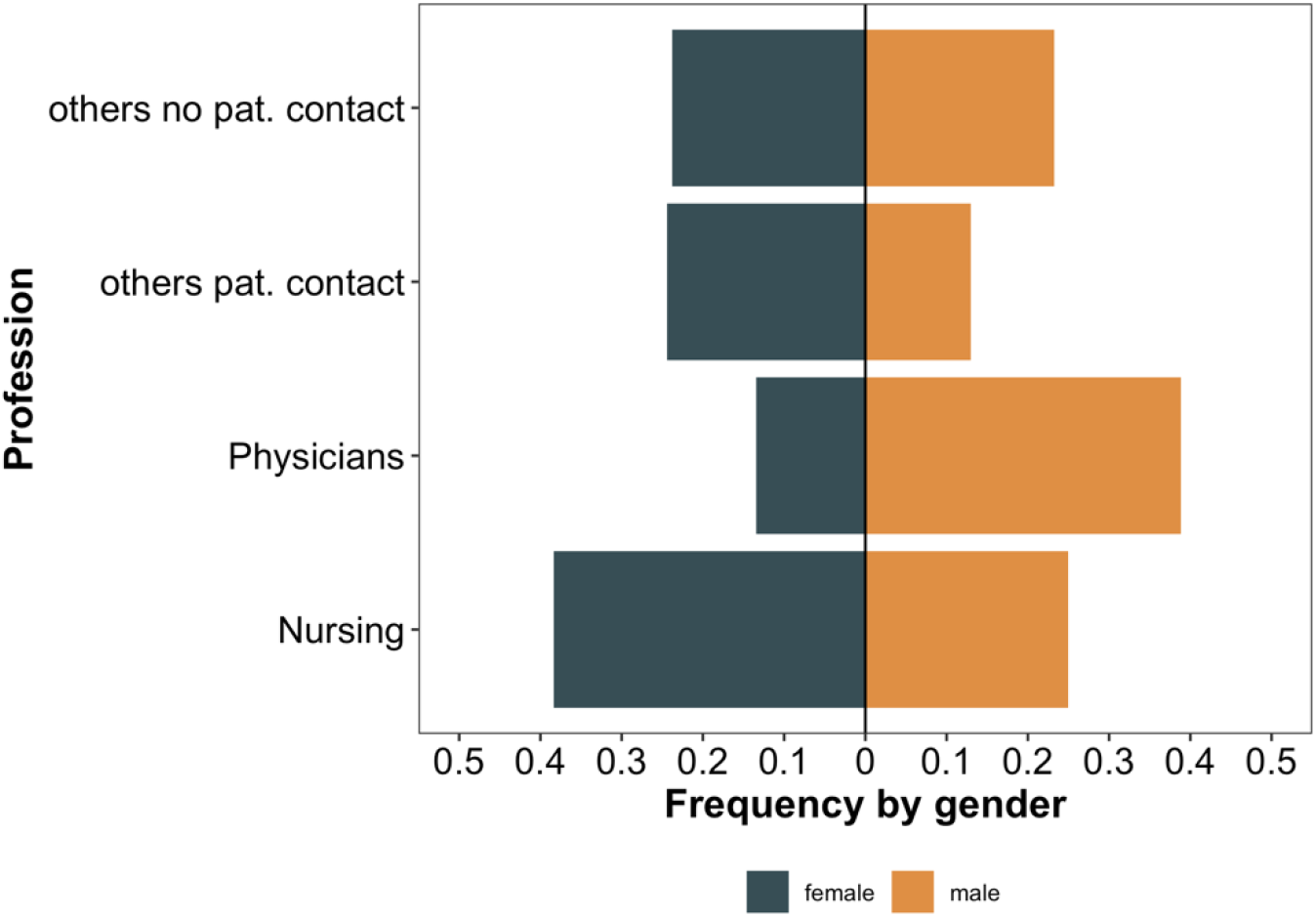
Gender separated distribution of HCWs’ profession. Relative frequencies of HCWs according to their profession and gender. Normalization of frequencies was performed for each gender separately.

**Supplementary Figure 2:**
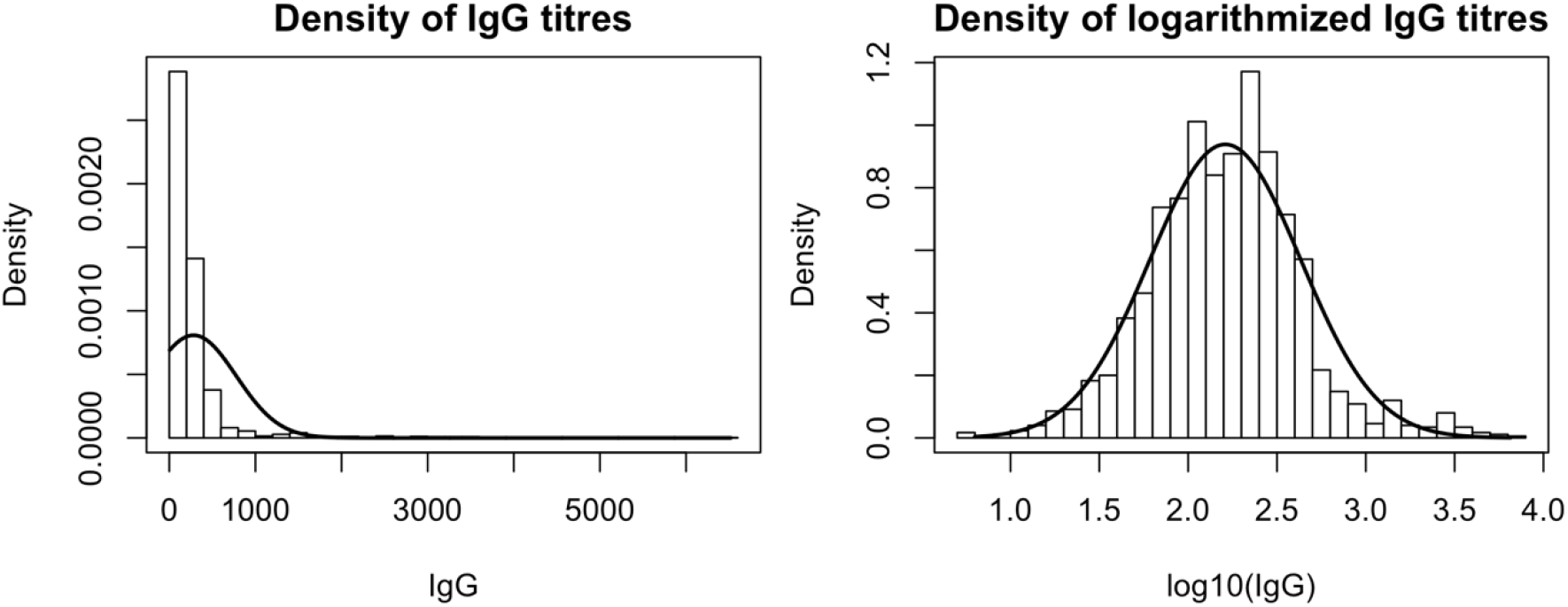
Distribution of Anti-SARS-CoV-2-Spike IgG levels. Density histogram depending on logarithmically scaled Anti-SARS-CoV-2-Spike IgG concentrations. Obtained logarithmized IgG levels of included study participants are nearly normal distributed.

**Supplementary Figure 3:**
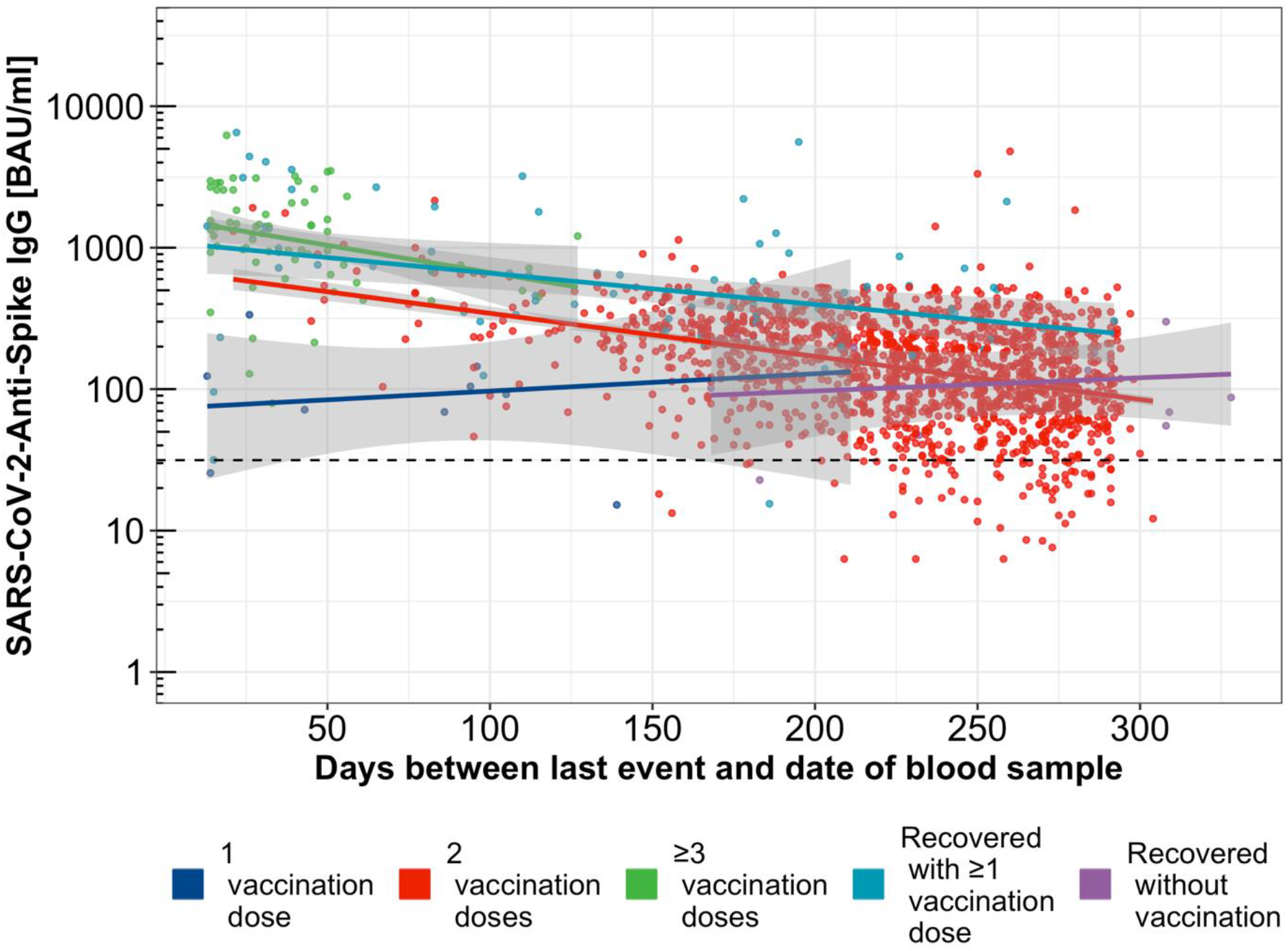
Temporal course of Anti-SARS-CoV-2-Spike IgG levels in the study subgroups. Temporal course of Anti-SARS-CoV-2-Spike IgG levels depending on time since last event for single (blue dots and regression line), double (red dots and regression line) and threefold or more (green dots and regression line) COVID-19 vaccinated as well as SARS-CoV-2 convalescent (purple dots and regression line) and hybrid immunized participants (turquoise dots and regression line).

**Supplementary Figure 4:**
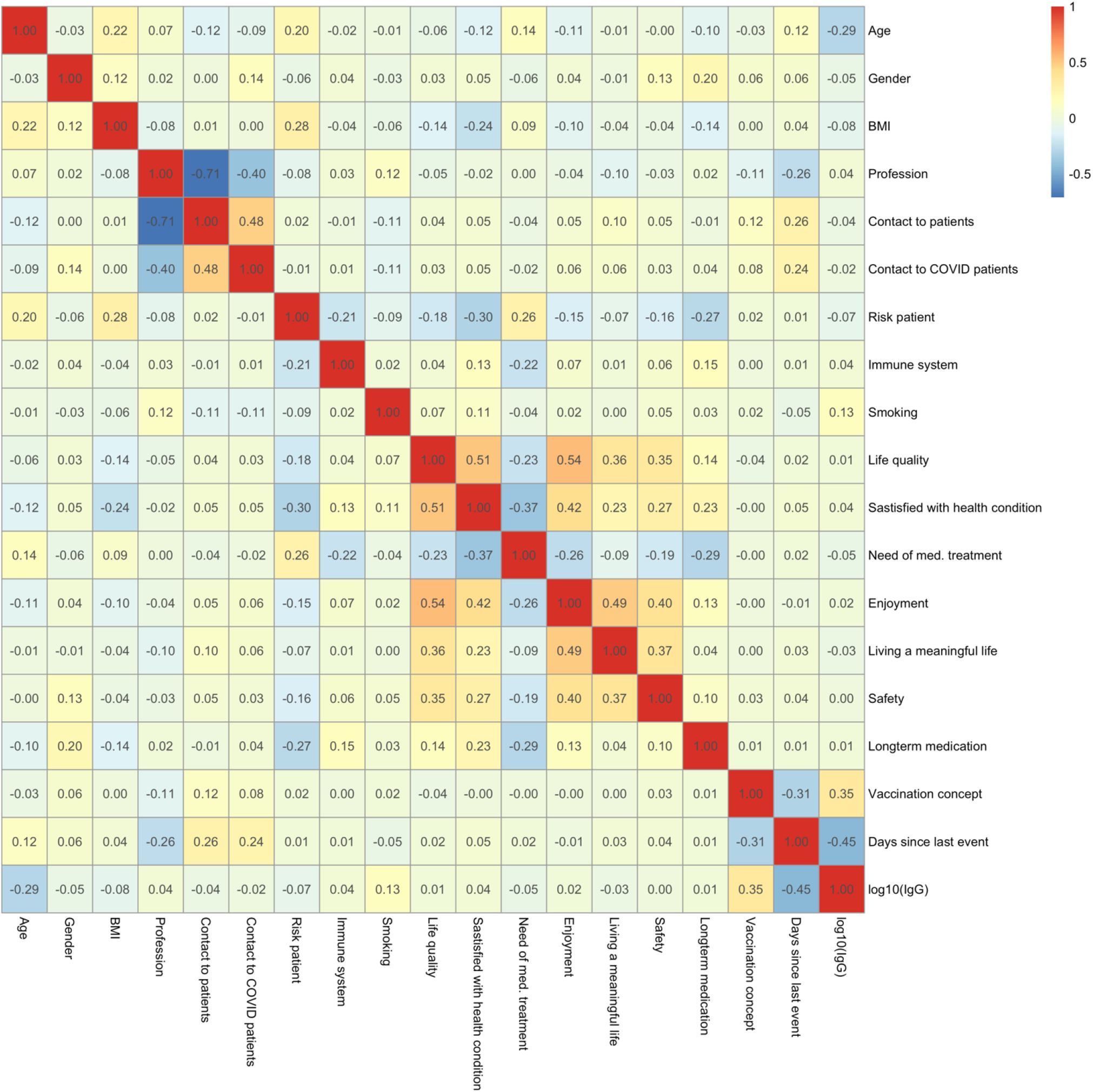
Correlation Anti-SARS-CoV-2-Spike-IgG titres and potentially associated factors. Heatmap of pairwise spearman correlation coefficients between Anti-SARS-CoV-2-Spike-IgG titres and all potentially influential factors. Colour scale orange to red indicates positive correlations, while colours from bright blue to dark blue indicate negative correlations.

**Supplementary Figure 5:**
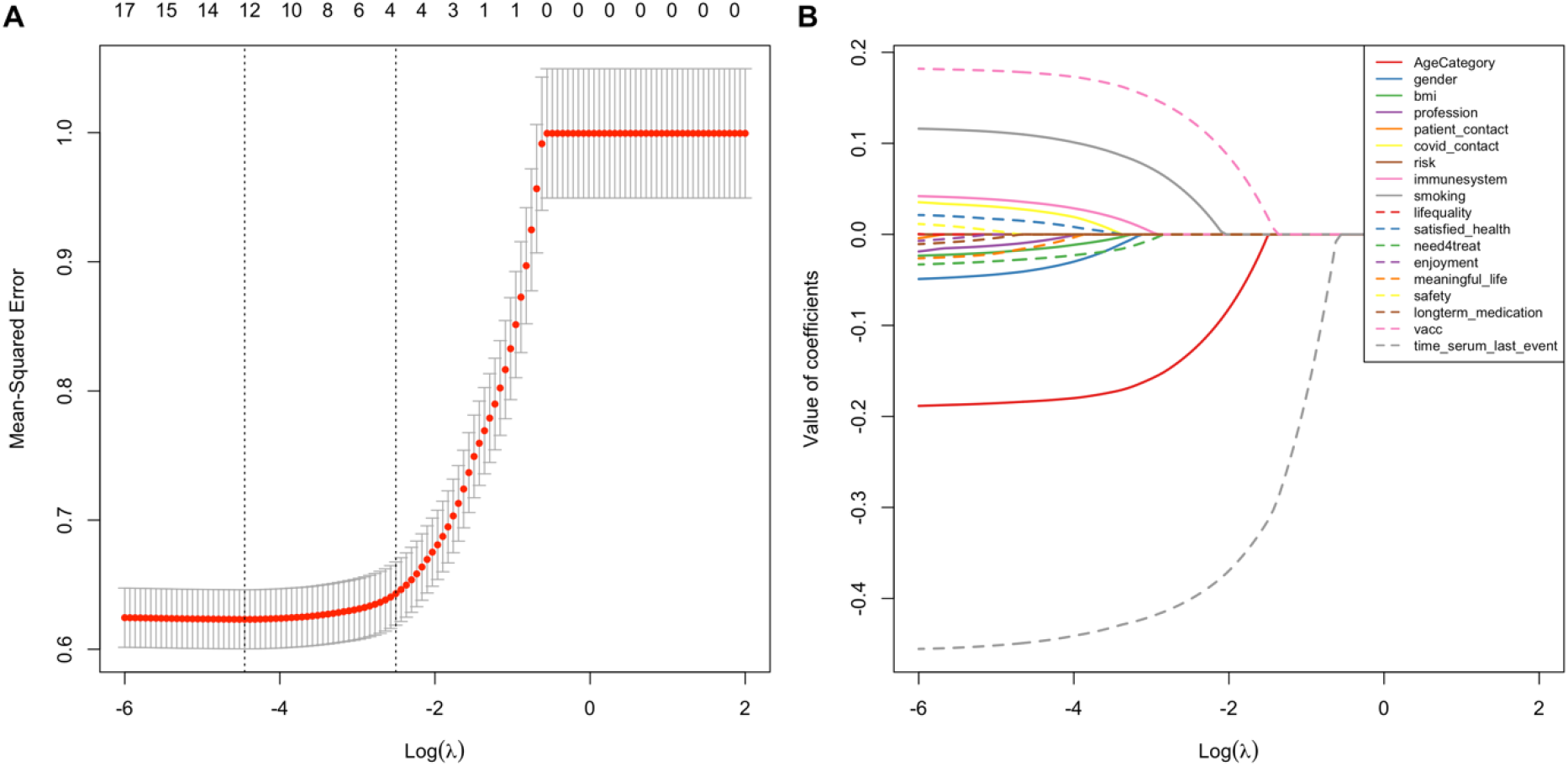
Cross-validation and factor selection of lasso regression model. Lasso regression for detection of factors associated to Anti-SARS-CoV-2-Spike-IgG titres. **5A**: Tenfold cross-validation procedure to determine optimal lambda parameter based on minimal mean-squared error. **5B**: Illustrating the shrinkage of coefficients (factors) towards zero with increasing lambda values.

**Supplementary Figure 6:**
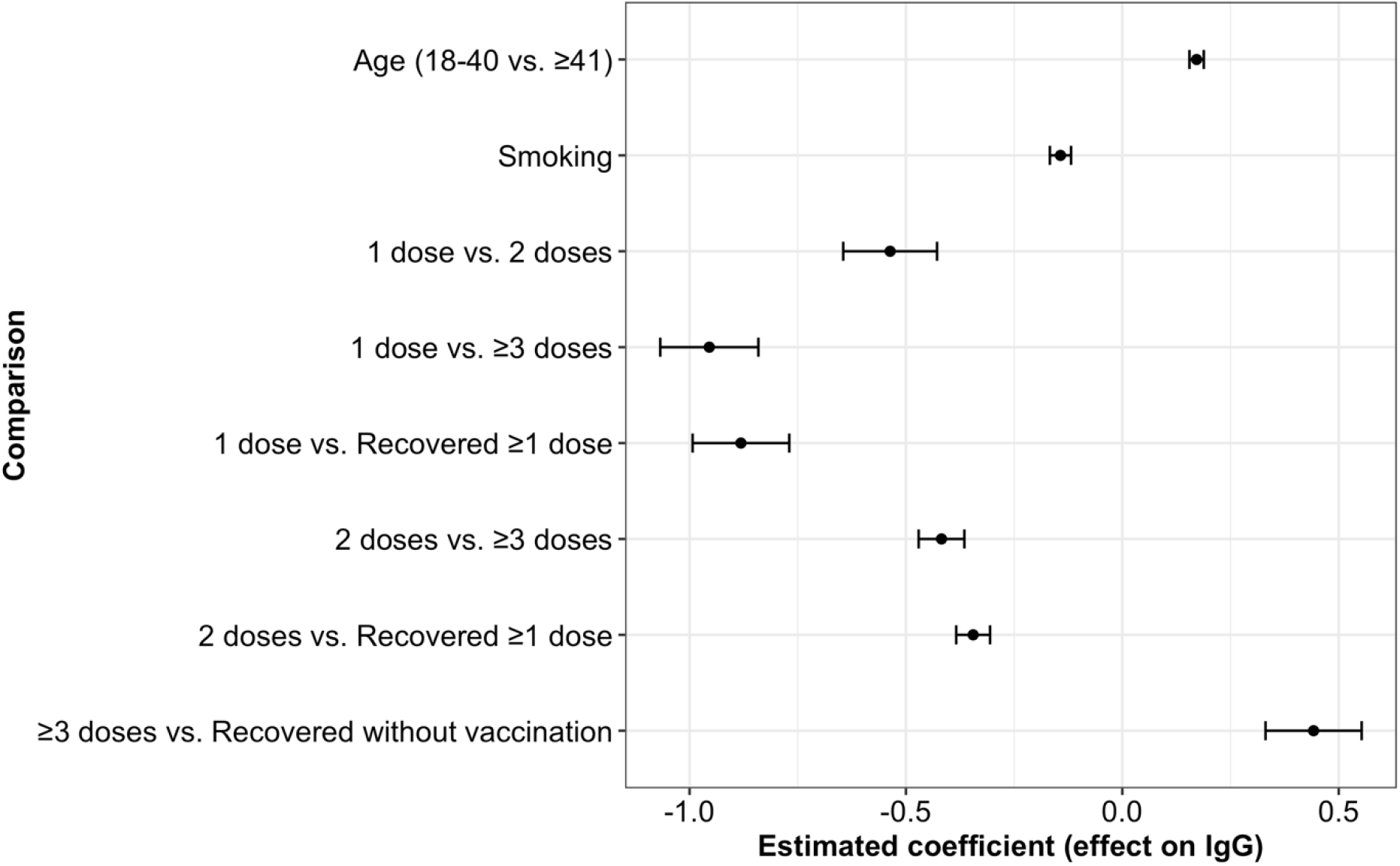
Post hoc analysis of pairwise differences. Results of post hoc analysis of pairwise comparisons. The abscissa shows the differences of the estimated marginal means while the ordinate shows all pairwise comparisons with statistically significant differences. The points represent the differences of the estimated marginal means, while the whiskers represent the estimated standard errors.

**Supplementary Figure 7:**
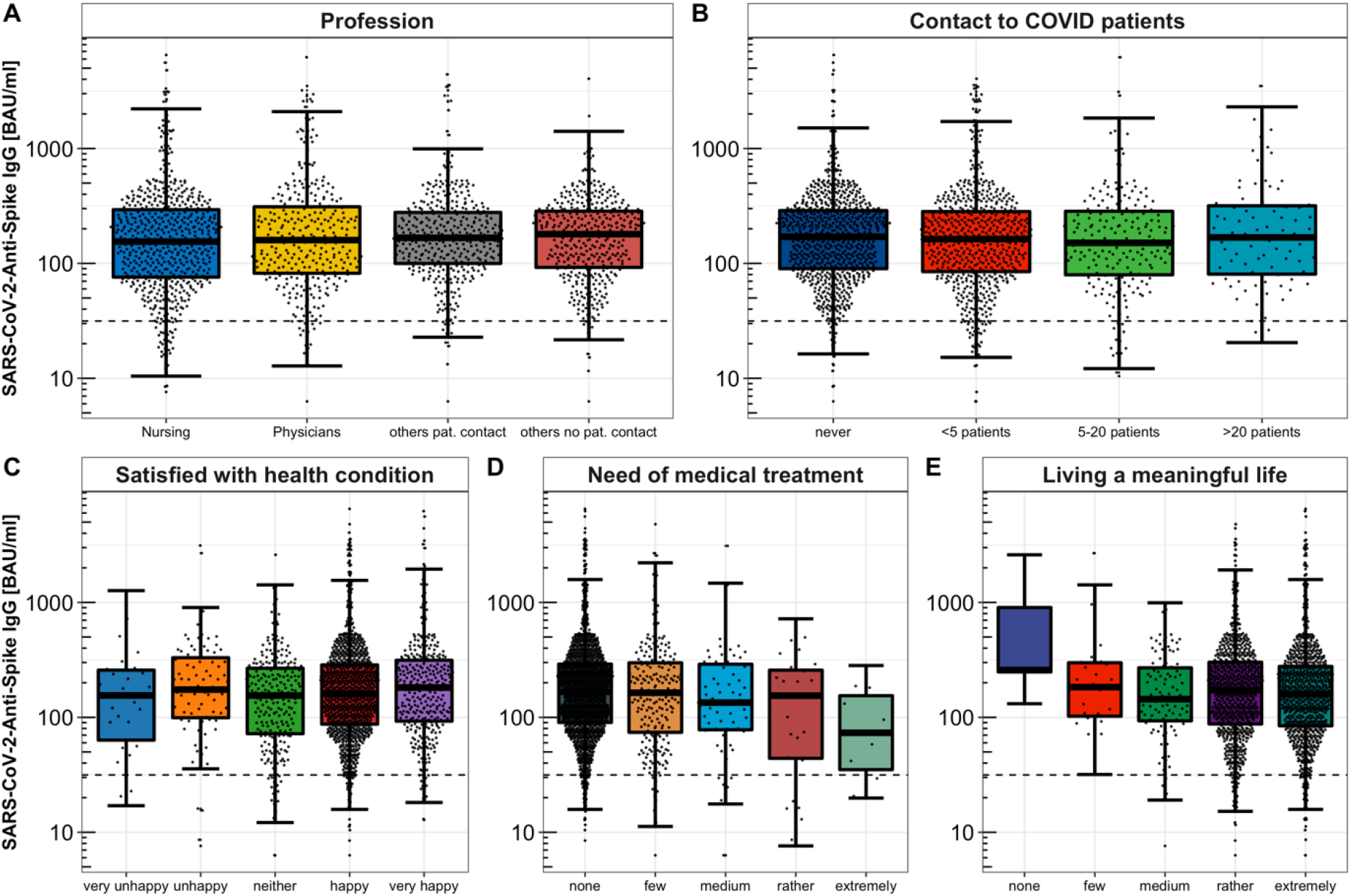
Comparison of Anti-SARS-CoV-2-Spike-IgG titres between groups of remaining factors. Pairwise comparisons of Anti-SARS-CoV-2-Spike-IgG titres between subgroups of remaining factors.

**Supplementary Table 1:**
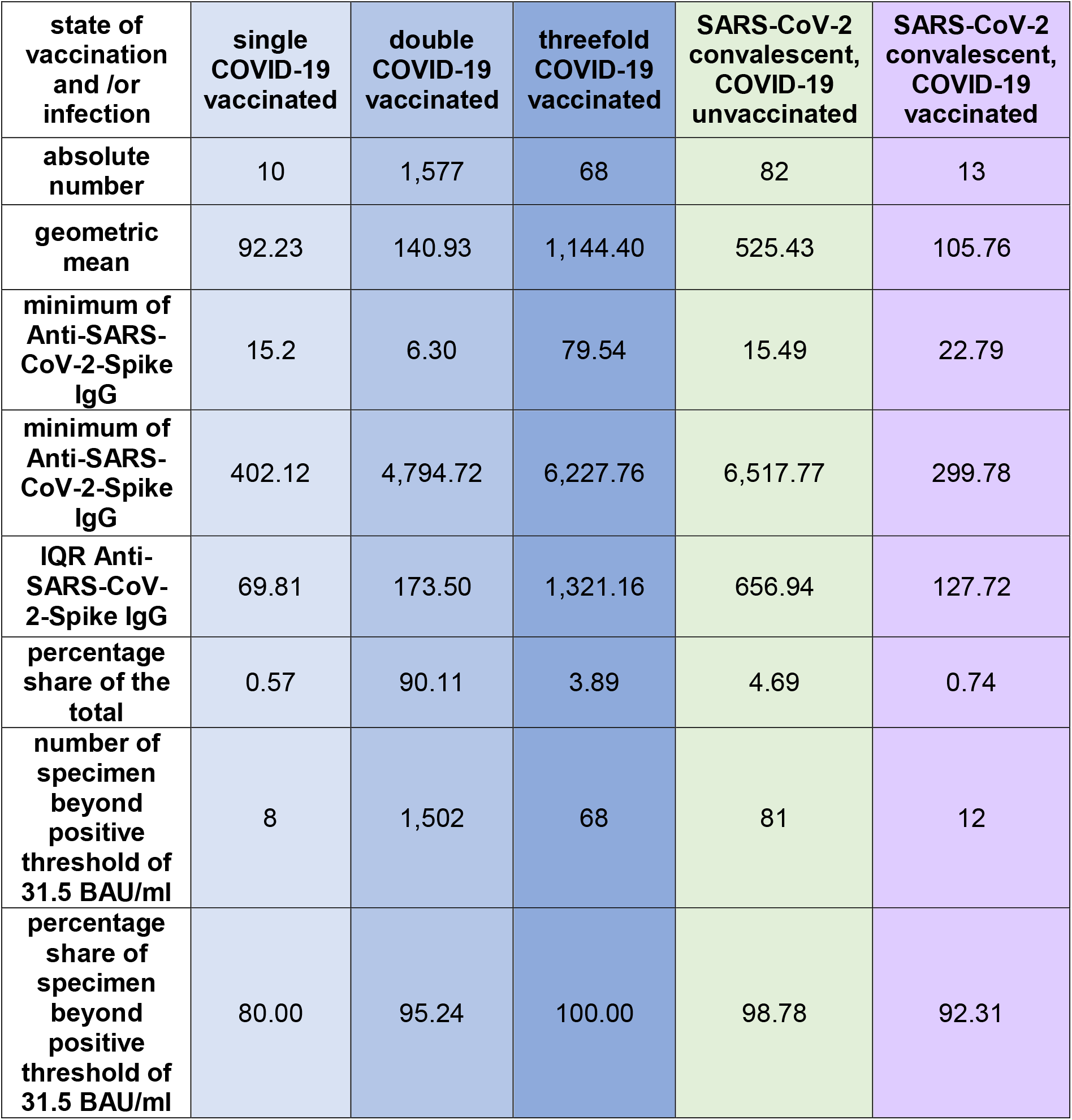

